# The role of spatial structure in the infection spread models: population density map of England example

**DOI:** 10.1101/2020.04.24.20077289

**Authors:** Gregory Mashanov, Alla Mashanova

**Affiliations:** The Francis Crick Institute, London, UK; The University of Hertfordshire, Hatfield, UK

## Abstract

In the current situation of a pandemic caused by COVID-19 developing models accurately predicting the dynamics of the outbreaks in time and space became extremely important.

Individual-based models (IBM) simulating the spread of infection in a population have a few advantages compared to classical equation-based approach. First, they use individuals as units, which represent the population, and reflect the local variations happening in real life. Second, the simplicity of modelling the interactions between the individuals, which may not be the case when using differential equations.

We propose to use freely available population density maps to simulate the infection spread in the human population on the scale of an individual country or a city. We explore the effect of social distancing and show that it can reduce the outbreak when applied before or during peak time, but it can also inflict the second wave when relaxed after the peak. This can be explained by a large proportion of susceptible individuals, even in the large cities, after the first wave.

The model can be adapted to any spatial scale from a single hospital to multiple countries.

## Introduction

In the last 30 years, the development of personal computers and high-level compilators (Kernighan & Ritchie, 1978) has allowed researchers to use Individual-Based Models (IBM) (see Gardner, 1970 for one of the first examples) to simulate complicated biological systems containing thousands of individuals (Mashanov, 1997). The use of object-oriented languages has allowed to code the properties of the objects of several classes and describe the corresponding functions generating object-specific events for the individuals interacting in a three-dimensional space (see Mashanov, 2014 for an example). It would be difficult to model spatially structured biological systems using classical mathematics (differential equations). One of the areas, where it became apparent, is epidemiology employing Susceptible-Infectious-Recovered (SIR) model https://en.wikipedia.org/wiki/Compartmental_models_in_epidemiology.

SIR models have been widely used to model infectious disease spread in epidemiology although some authors question the validity of these models’ predictions because of rather unrealistic assumptions (e.g. Huppert & Katriel, 2013). One of the common assumptions of SIR models is that populations are well-mixed. This means that any individual is equally likely to come into contact with any other individual ignoring the fact that people living in large cities are much more likely to come into contact with each other. Modelling spatial structure explicitly allows overcoming this problem.

Here, we propose to use Population Density Maps (PDM) to get a realistic distribution of the population on a scale of a country or a smaller region. We used the simplest IBM with a minimal number of properties, parameters, and functions to show the advantages of PDM-based IBM in understanding the temporal and spatial dynamics of an epidemics in a real human population.

### Model Description

We constructed a basic IBM, where every individual has the following properties: floating-point X and Y coordinates, and integer “Infection status”, and “Density index” (Table 1).

**Table 1.** Properties of individuals

| Name | Explanation | Variable type |
| --- | --- | --- |
| Position | X and Y coordinates defining position on the map | Floating-point |
| Infection status | "Susceptible" or "infectious". Individual becomes "recovered" after a certain number of days (defined by "Duration of infection") | Integer (0 – susceptible, above 0 - days since infection) |
| Density index | When PDM is used, an individual is assigned to pixel values with a certain population density. When moving or commuting the individuals remain within the same density | Integer (1 in "no spatial structure", 1 to 255 in PDM) |

The list of parameters is given in Table 2 and the list of functions in Table 3. At the start (“NewRun()”), individuals are placed randomly on the plain map, randomly inside the shape map, or randomly, but proportionally to PDM values, when PDM is loaded. Events happening at every step (equal to one day) are described in the “Cycle()” function. At every step, every individual can move (“Move()”) and a fraction of individuals (chosen randomly) is allowed to commute (“Commuting()”). Individuals move randomly within the “Mobility” distance. The new position should belong to a pixel marked as “land” for the model with no spatial structure or, in case of PDM, or to a pixel with the same density value, as the pixel were the object was placed at the beginning of the run (Table 1). This restriction allowed the individuals to move within populated areas, but without spreading across the whole map. It was also important to use “Commuting()” function to allow some individuals to move long distances. Commuting is also confined to the pixels with the same density values – so that commuters from large cities go to another large city. The fraction of commuters could be set to zero to run the model in the “local spread” mode.

**Table 2.**
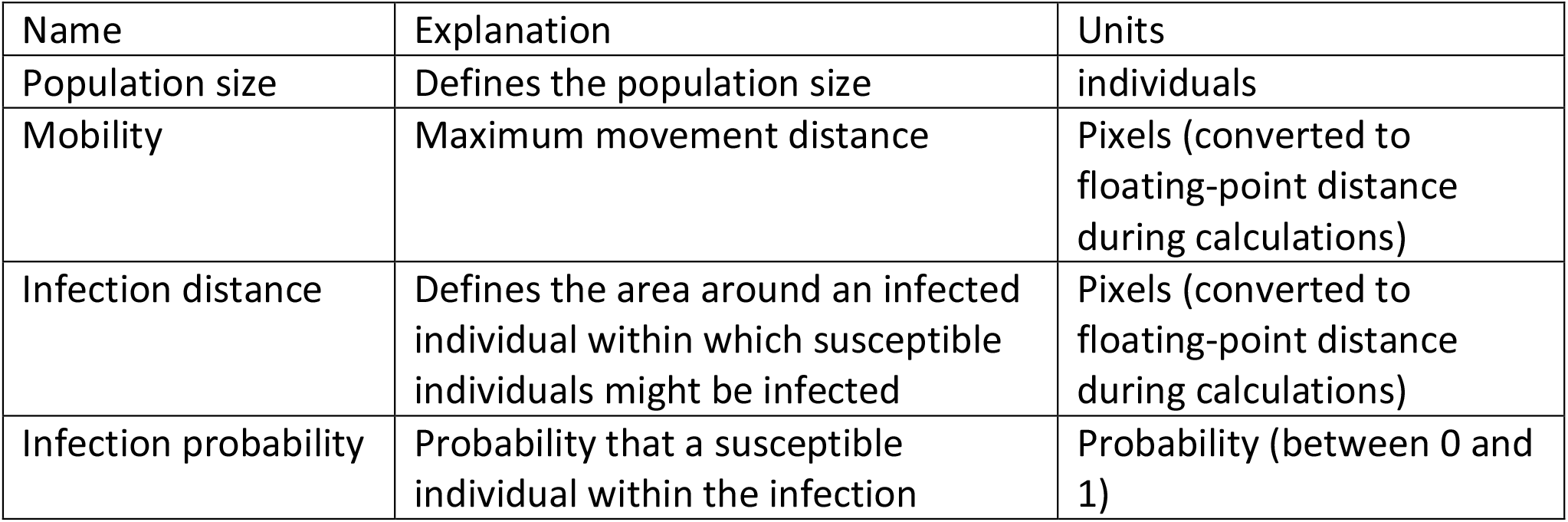
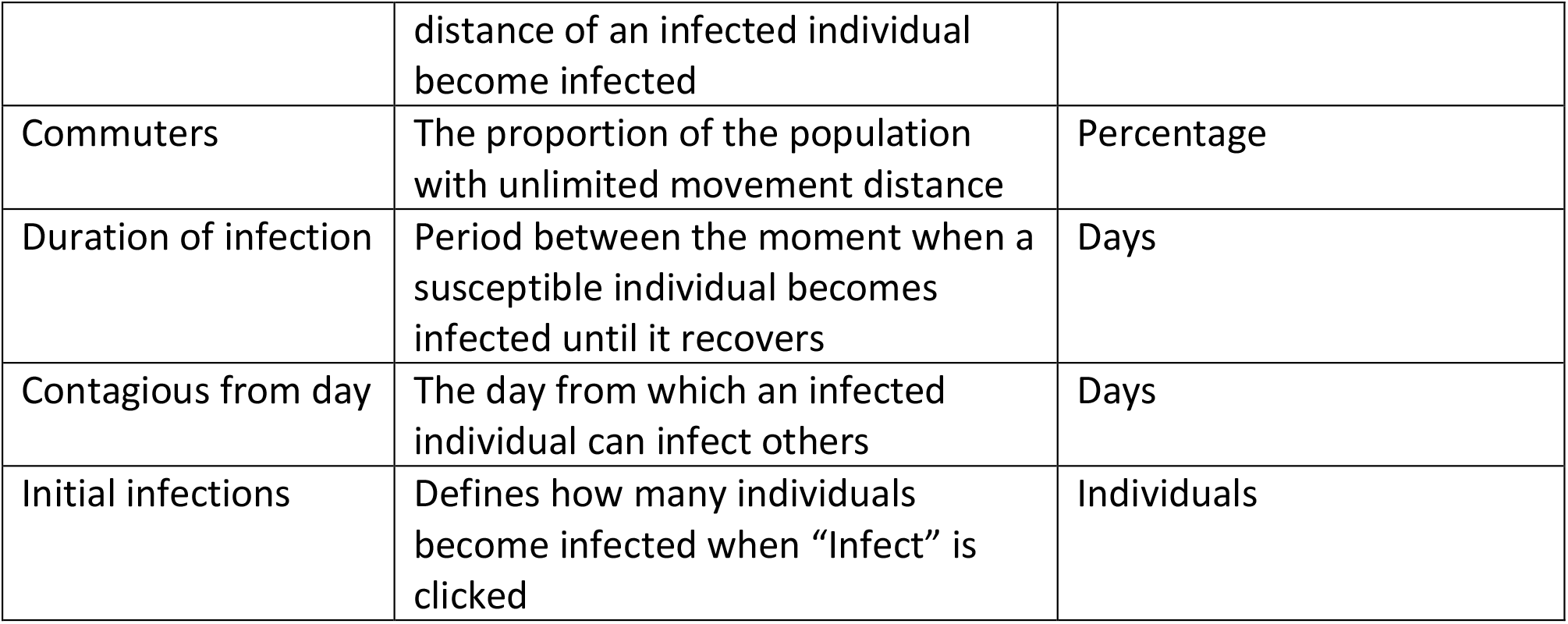
Parameters and variables (Population tab)

**Table 3.**
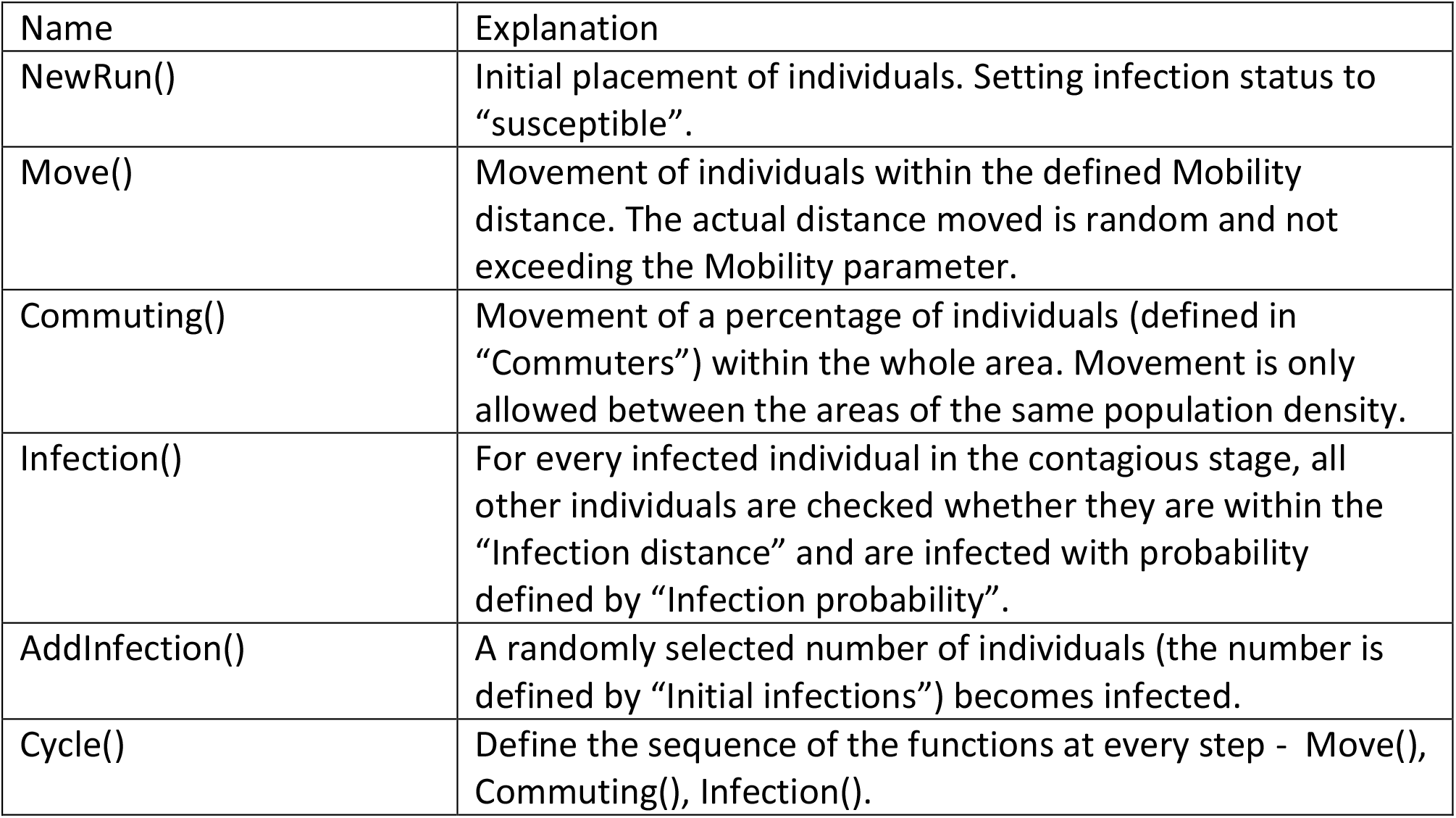
Functions used in the model (see Appendix for codes)

| Name | Explanation |
| --- | --- |
| NewRun() | Initial placement of individuals. Setting infection status to “susceptible”. |
| Move() | Movement of individuals within the defined Mobility distance. The actual distance moved is random and not exceeding the Mobility parameter. |
| Commuting() | Movement of a percentage of individuals (defined in “Commuters”) within the whole area. Movement is only allowed between the areas of the same population density. |
| Infection() | For every infected individual in the contagious stage, all other individuals are checked whether they are within the “Infection distance” and are infected with probability defined by “Infection probability”. |
| AddInfection() | A randomly selected number of individuals (the number is defined by “Initial infections”) becomes infected. |
| Cycle() | Define the sequence of the functions at every step - Move(), Commuting(), Infection(). |

Infections can be introduced at any moment during the model run by clicking “Infect” which runs “AddInfection()” function. Specified by “Initial infections” number of individuals are selected randomly from the population and become infected. “Contagious from day” parameter defines when the infected individuals can infect others. For the individuals in the infectious state (time interval between “contagiousFrom” and “contagiousTo”), the “Infection()” function was used to check for other susceptible individuals within the “Infection Distance”. A random number generator was used to determine if these individuals became infected according to the “Infection Probability”.

The model is written in C language using C++ Builder XE7 (Embarcadero, CA). The code used to display the results and set the parameters was omitted from the code in the Appendix. Otherwise, this is the whole description of the model used to produce all the data in this publication. The model is available to download from Zenodo DOI:10.5281/zenodo.3763585 or www.mashanov.uk.

## Results

We first tested our model without introducing spatial structure (Fig. 1). In all examples shown in Fig. 1, the average population density was kept 0.1 individual/pixel, mobility 2 pixels, infection probability 0.02, infectious period 20 days, the individual is contagious from day 2 (Table 4). Fig. 1A and movie 1 show an infection spread in the “plane field” with no commuters. In these conditions, we failed to reproduce any spread of the infection with the infection distance less than 4 pixels. Without commuting, the spread was very slow with less than 2% of the population infected at any time. If we introduce “commuting” (3%), the spread accelerates reaching ∼30% of the infectious individuals at the peak (Fig. 1B). But the epidemic would not start if we reduce the infection distance < 3 pixels (data not shown). When we introduced a shape map of England and Wales (Fig. 1C) with the same population density of 0.1 individual/pixel and other conditions as in Fig. 1B, the results of the model run were very similar to the results from the “plane map” run.

**Table 4.** Parameters for the examples shown in figures 1 and 2. In all examples, initial infectious were introduced at the beginning.

| Parameter | 1a | 1b | 1c | 2a | 2b | 2c | 2d |
| --- | --- | --- | --- | --- | --- | --- | --- |
| Population size | 10000 | 10000 | 35000 | 41500 | 41500 | 41500 | 41500 |
| Mobility | 2 | 2 | 2 | 2 | 2 | 2 | 2 |
| Infection distance | 4 | 4 | 4 | 4 | 2 | 1.5 | 4 |
| Infection probability | 0.02 | 0.02 | 0.02 | 0.02 | 0.02 | 0.02 | 0.01 |
| Commuters | 0 | 3 | 3 | 3 | 3 | 3 | 3 |
| Duration of infection | 20 | 20 | 20 | 20 | 20 | 20 | 20 |
| Contagious from day | 2 | 2 | 2 | 2 | 2 | 2 | 2 |
| Initial infections | 2 | 2 | 2 | 2 | 2 | 2 | 2 |
| PDM used | no | no | no | yes | yes | yes | yes |

**Fig. 1.**
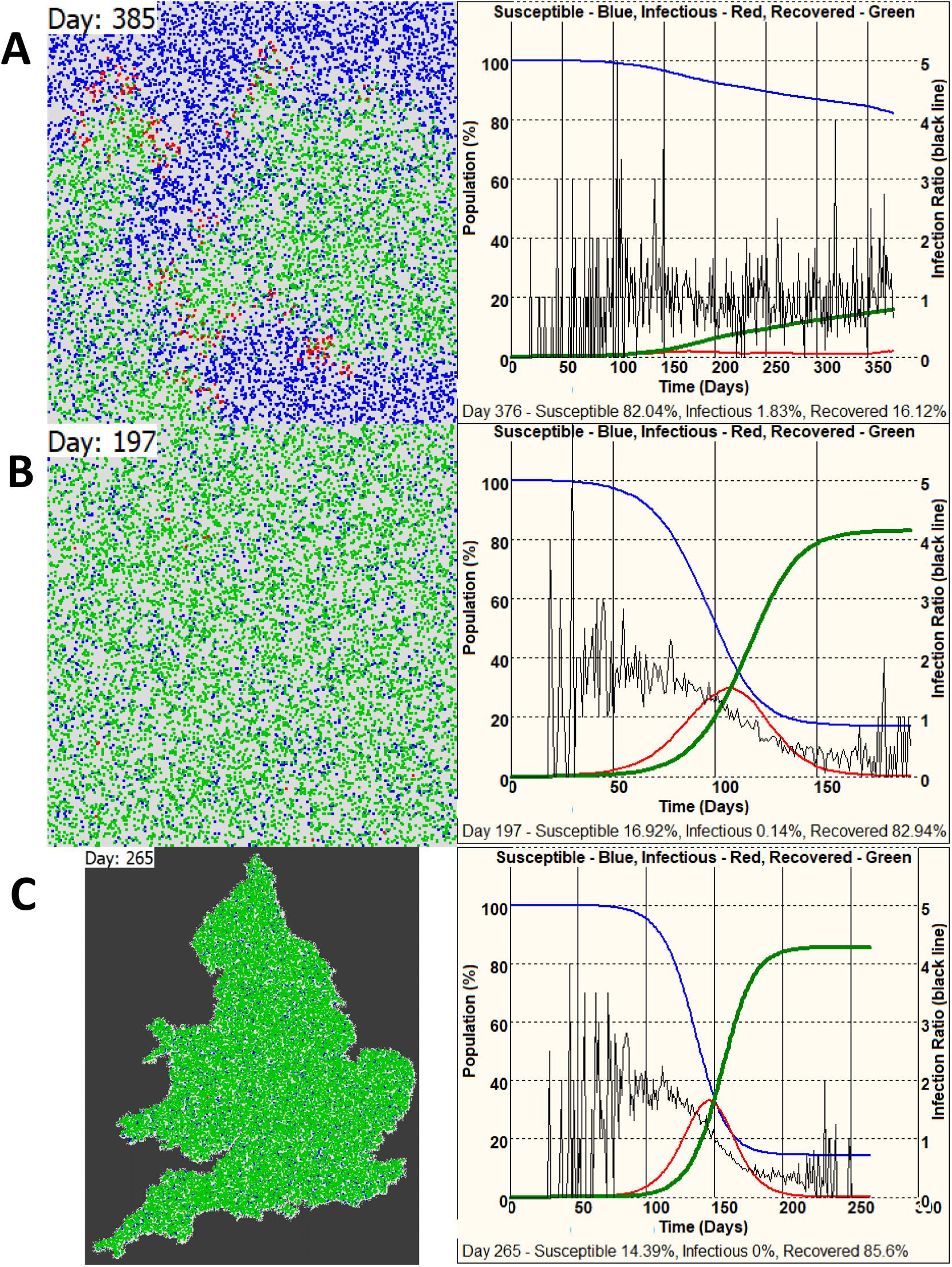
The infection spread without spatial structure. **A** – Individuals are placed on a square map, where they are allowed to move and to infect other individuals (see text for the description of the model). The map on the left shows the distribution of individuals: blue – susceptible, red – infectious, green – recovered (the same colours are used in all other figures). The graph on the right shows the population dynamics (colours as on the map). The black line (right Y-axis) shows the average number of individuals infected by a single infected individual during the infectious period. **B** – The same as A, but with commuting (3% of the population). **C** – England and Wales boundary map was used. Population size was adjusted to keep the density the same as in B. Other conditions are the same as in B (see text and the tables for the full description of the conditions).

We introduced spatial structure using PDM of England (Census 2011, https://en.wikipedia.org/wiki/Demography_of_England), where the map pixel values ranged from 1 to 255 (8-bit greyscale). At the start, the individuals were placed randomly but proportionally to the map pixel values. When we run our model using the same average population density 0.1 individual/pixel and parameters as in Fig. 1BC, we observed a sharp rise of infection reaching the peak of ∼50% of infectious individuals (∼80% recovered at the end of the epidemic, Fig. 2A, movie 2). The significant rise in infection can be explained by the fact that, in some areas (e.g., in the London area), the population density reached ∼40 individuals/pixel. If we reduce the infection distance by half (2 pixels) the peak of infection is reduced to ∼15% of infectious individuals (Fig. 2B). Further decrease of this value to 1.5 pixels brings the peak of infection to ∼3.5% (∼25% of recovered individuals, Fig. 2C). If instead of reducing infection distance from 4 to 2 pixel, we reduce the probability of infecting from 0.02 to 0.01 we observe the reduction in the peak of infection from ∼50% to ∼30% (See Fig. 2B and 2D for comparison).

**Fig. 2.**
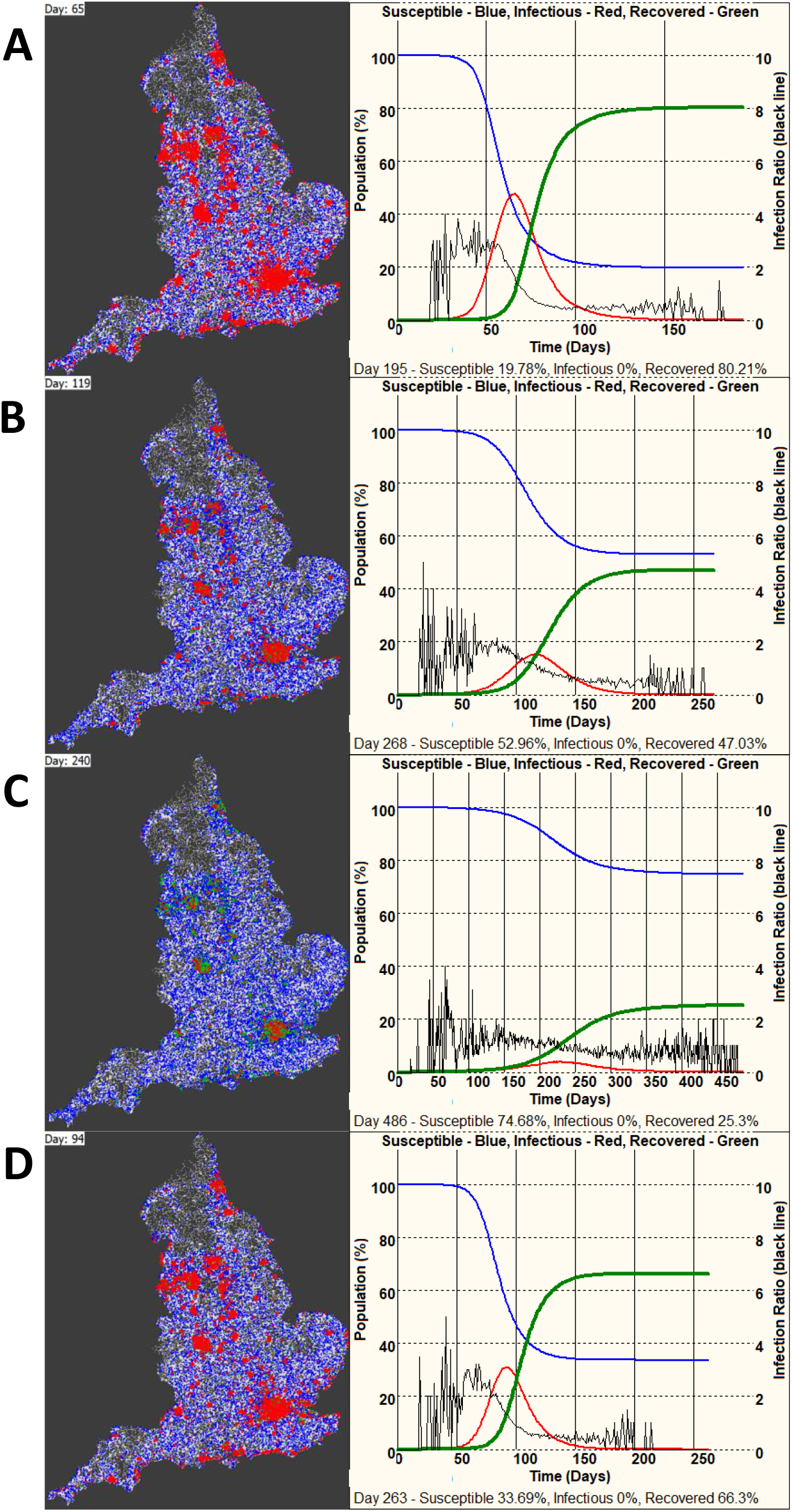
The infection spread with spatial structure (Population Density Map of England). **A** – Conditions as in Fig. 1C. Population size was adjusted to keep the density as in Fig. 1. The map on the left shows the distribution of susceptible (blue), infectious (red), and recovering (green) individuals at the peak of the epidemic. The graph on the right shows population dynamics (colours as on the map). The black line (right Y-axis) shows the average number of individuals infected by a single infected individual during the infectious period. **B** - the same conditions as in A, but the infection distance was reduced from 4 to 2 pixels. **C** - the same conditions as in A, but the infection distance was reduced to 1.5 pixels. **D** - the same conditions as in A, but the infection probability was reduced from 0.02 to 0.01. NOTE – the graphs show the population dynamics across the whole country. These values vary regionally.

We explored the effect of the reduction of infection distance, which can be interpreted as social distancing, during an outbreak. Introducing social distancing, in the form of reducing the infection distance from 4 to 1.5 pixels, in the middle of an outbreak, which would reach ∼50% of infectious individuals at its peak, resulted in quick slowing down of the outbreak, reaching the plateau at ∼32% (Fig. 3A). However, introducing social distancing before outbreak leaves a large fraction of the population susceptible. If we remove social distancing following the end of the first weak outbreak (increase the infection distance from 1.5 to 4 pixels) we can inflict a second outbreak with around 25% of infected individuals at its peak, by introducing just two infected individuals into the population (Fig. 3B, movie 3).

**Fig. 3.**
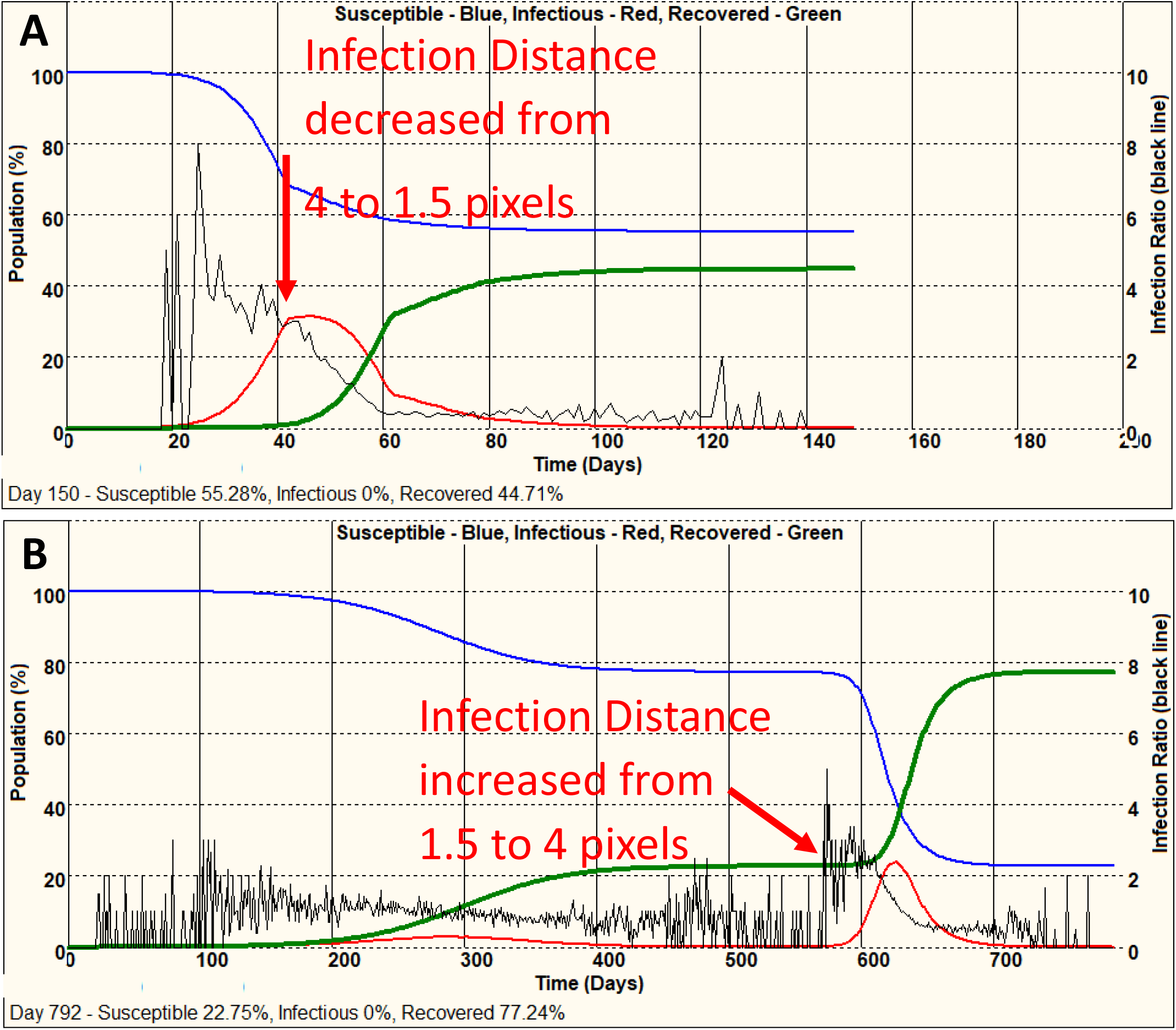
The effects of changes in social distancing during and after an outbreak. **A** – social distancing was introduced in the middle of an outbreak, which would reach 50% of infectious individuals at its peak. This measure resulted in quick slowing down of the outbreak, reaching the plateau at ∼32%. **B** – If we run the model with severely restricted infection distance (1.5 pixels, See Fig. 2C), we will observe a very mild epidemic, reaching ∼3.5% of the infectious individuals at its peak. However, if we relax the social distancing by increasing infection distance to 4 pixels, we will observe the second outbreak with ∼25% of infected individuals at its peak.

The secondary axis in all figures shows “Infection ratio” curve, which is the average number of individuals infected by a single infected individual during the infectious period. Since it represents the interaction between the basic reproductive number R0 and the proportion of susceptible individuals in the population, in all graphs it decreases as the number of susceptible individuals decrease, and this decrease precedes the peak/decrease of the outbreak.

## Discussion

In this work, we presented a simple individual-based model, which has around 100 lines of code and 5 functions describing interactions between individuals. The model accurately reproduces the dynamics of classical susceptible-infectious-recovered (SIR) models (see Fig. 1 for example). It was important to introduce the “Commuting()” function into our model, otherwise, the spread of infection from a few individuals was very slow (movie 1), even if large infection distance or infection probability values were used.

It is generally accepted that the population density greatly affects the dynamics of infection spread either in human or, in fact, in many animal communities (e.g. Tildesley et al. 2010). Population density is often uneven, especially in case of developed human society, where the majority of the population is concentrated in large settlements (i.e., towns and so on). This leads to variation between different regions (Merler & Ajelli 2010, Brizuela et al. 2020). To take into account this critically important parameter, we introduced Population Density Map concept into our model: the probability of placing individuals at the beginning of a run depends on PDM values, and the movements are restricted to the pixels with the same population density. The use of PDM made our model much more realistic (See Fig. 2 and movie 2). Running the model using PDM with the same average density (0.1 individual/pixel) and other parameters, as in the case of a plane map, we observed the very fast epidemics affecting 50% of the population in 50 days since the start (Fig. 2A) as opposed to about 30% in 150 days (Fig. 1C).

We also have found that the infection distance, which can be modified in real life by social distancing, severely affects the spread of infection: reducing the infection distance from 4 to 2 and 1.5 pixels with all the other parameters kept constant, reduced the peak of infection from ∼50% of infected individuals to 15% and 3.5% correspondingly. If we reduce this distance further to 1 pixel, the epidemic fails to start at all, even in the situation when the initial infection starts in a highly-populated area (for example, in London area). However, the model predicts that relaxing social distancing after the first peak led to an even higher secondary outbreak started by introducing only 2 new infected individuals. The probability of the second outbreak was higher when some big cities escaped the first outbreak.

Dieckmann et al. (2020) reviewed the potential of simulations for the variety of uses in the healthcare organisations, including education and optimisation of work structure in the light of COVID-19 crisis. Our paper aims to advertise the use of IBM models because they are easy to modify and can be applied to different situations. The infection spread IBM, presented here can be easily expanded to include many important parameters of individuals in real human population: age, age-related population distribution and mortality, ethnic differences, and many others.

## Conclusion

We propose a very simple IBM using population density maps to simulate the infection spread (for example COVID19) in real situations. Any existing PDM, on a scale of a country or a smaller region, can be used to explore the temporal and, most importantly, spatial dynamics of the epidemics to predict and manage the future infection outbreaks.

## Data Availability

The executable version of the described model is available for download from Zenodo DOI:10.5281/zenodo.3763585 or www.mashanov.uk

http://www.mashanov.uk

DOI:10.5281/zenodo.3763585

## Acknowledgements

We would like to thank Dr Justin Molloy (the Francis Crick Institute) for encouraging this project. This work was supported by the University of Hertfordshire and by the Francis Crick Institute which receives its core funding from Cancer Research UK (FC001119), the UK Medical Research Council (FC001119), and the Wellcome Trust (FC001119).

## Supplementary materials

1. Appendix (see below). The model code.
2. Movie 1 – Slow spread of the infection on the plane map in the absence of commuting.
3. Movie 2 – Fast spread of infection when using population density map (England example).
4. Movie 3 – The two infection outbreaks. The second, much bigger, epidemics was caused by the increase in infection distance from 1.5 to 4 pixels.

## Appendix

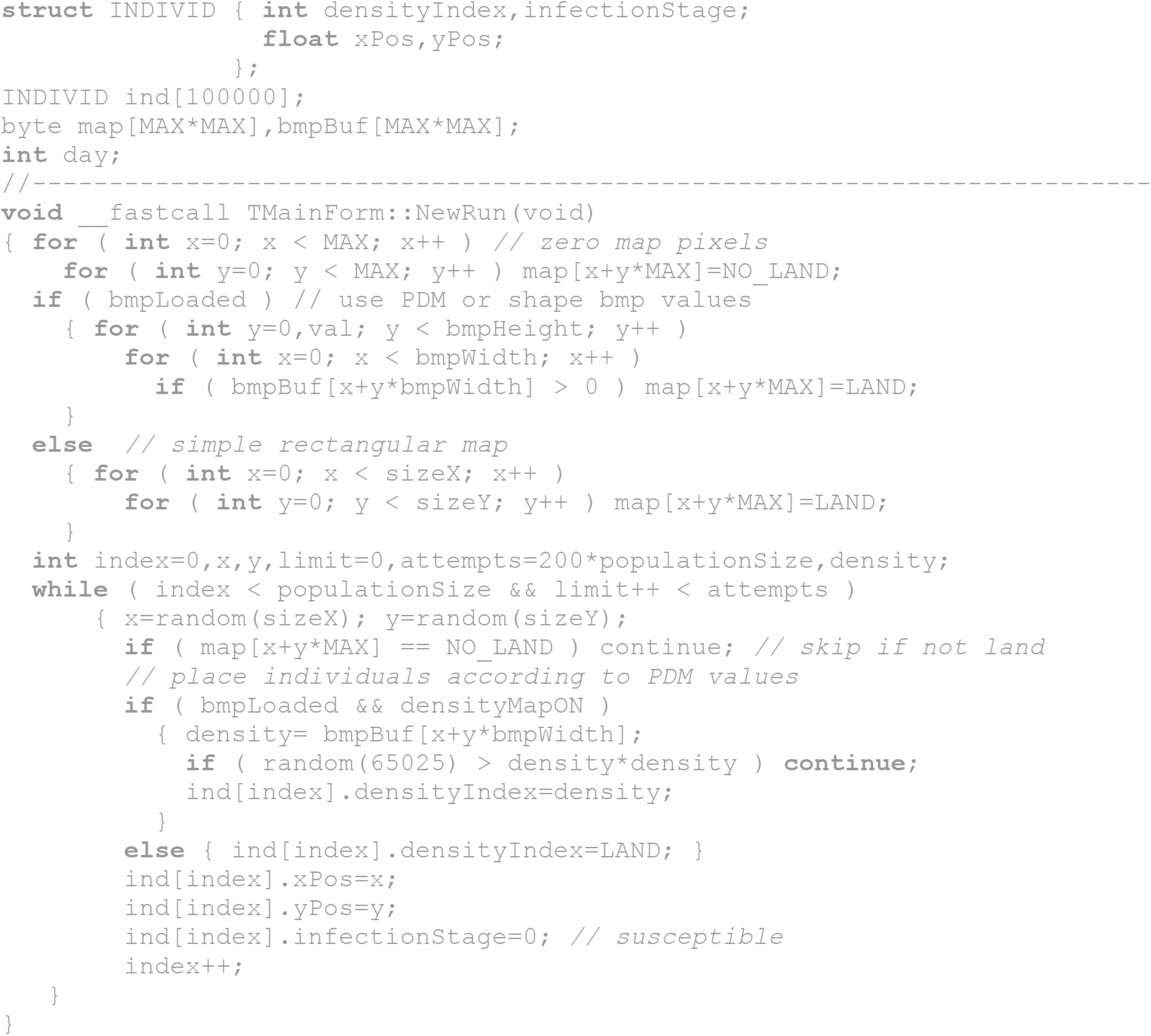

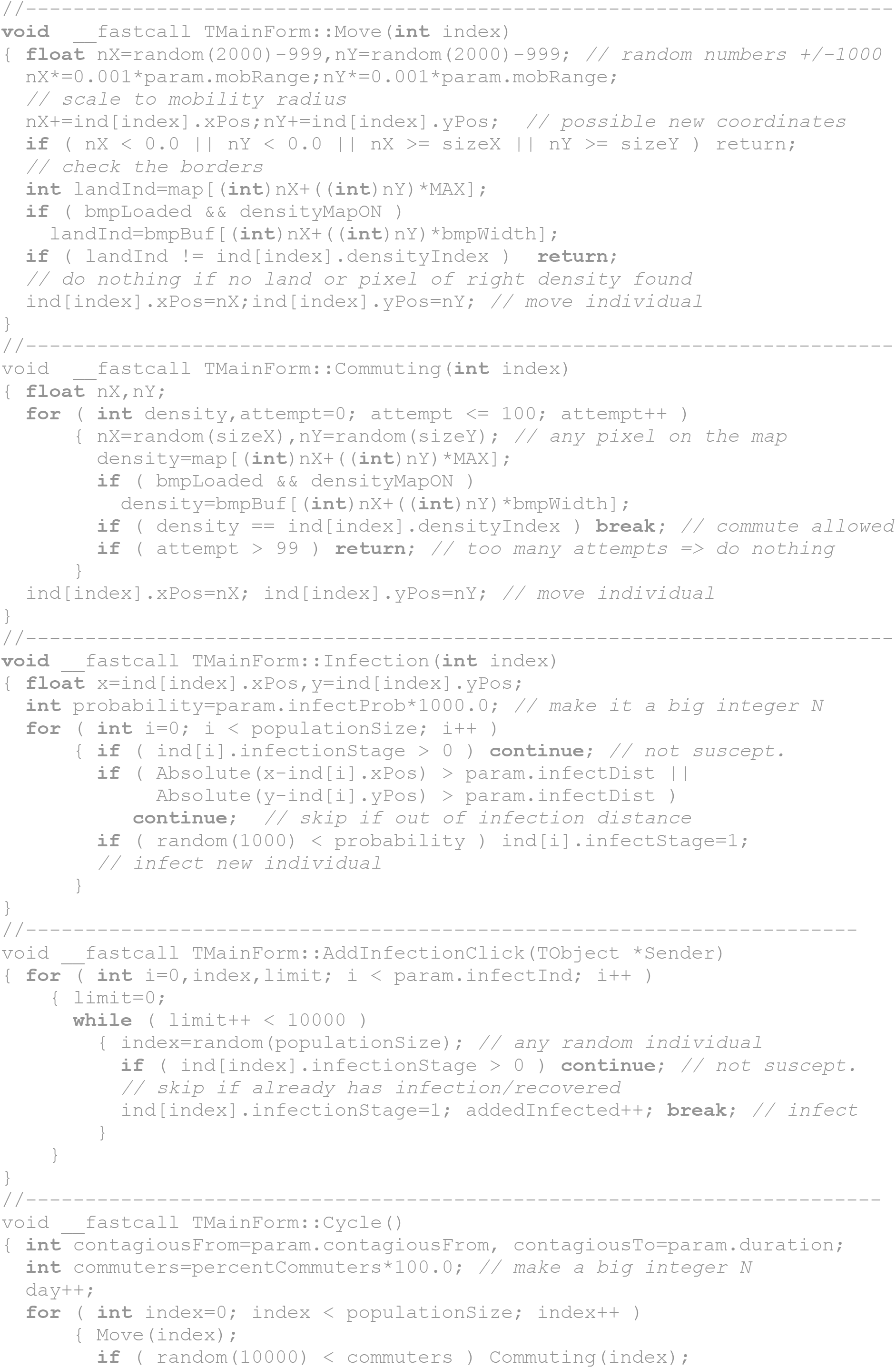

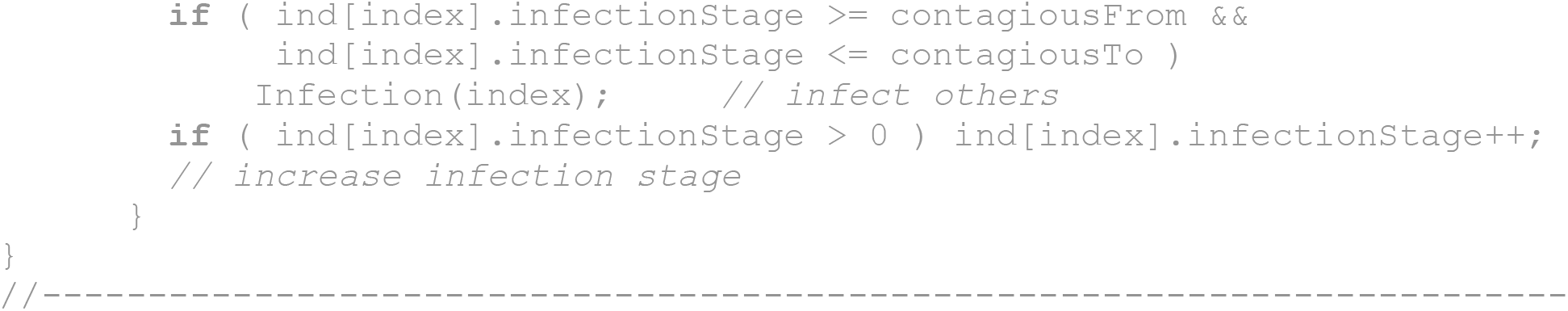

## References

Dieckmann P et al. 2020. The use of simulation to prepare and improve responses to infectious disease outbreaks like COVID-19: practical tips and resources from Norway, Denmark, and the UK. Advances in Simulation 5:3 https://doi.org/10.1186/s41077-020-00121-5.

Gardner M. 1970. Mathematical Games - The Fantastic Combinations of John Conway’s New Solitaire Game ‘Life’, Scientific American 223: 120–123.

Huppert A, Katriel G. 2013. Mathematical modelling and prediction in infectious disease epidemiology. Clin. Microbiol. Infect. 19: 999–1005.

Kernighan BW, Ritchie DM. 1978. The C Programming Language (1st ed.). Englewood Cliffs, NJ: Prentice Hall. ISBN 0-13-110163-3.

Mashanov GI. 1997. A cross-bridge model for the artificial mobile systems. Biofizika 42(5):1113–21.

Mashanov GI. 2014. Single molecule dynamics in a virtual cell: a three-dimensional model that produces simulated fluorescence video-imaging data. J R Soc Interface. 11(98): 20140442.

Merler S., Ajelli M. 2010. The role of population heterogeneity and human mobility in the spread of pandemic influenza. Proceedings. Biological sciences, 277(1681), 557–565. https://doi.org/10.1098/rspb.2009.1605

Tildesley MJ et al. 2010. Impact of spatial clustering on disease transmission and optimal control. PNAS 107(3):1041–1046. https://doi.org/10.1073/pnas.0909047107

